# Beyond Billing Codes: An Agentic Large Language Model Pipeline for Extracting Glaucoma Procedural Data from Free-Text Clinical Notes

**DOI:** 10.64898/2026.10.29.26366154

**Authors:** Janet Kang, Nicholas Solages, Gustavo A. Samico, Naomi E. Gutkind, Rafael Scherer, Felipe A. Medeiros, Swarup S. Swaminathan

## Abstract

**Purpose:** To evaluate the accuracy of a large language model (LLM)-based agentic workflow to extract procedural data from free-text glaucoma clinical notes in the electronic health record (EHR).

**Design:** Cross-sectional.

**Subjects:** 1,250 subjects in the Bascom Palmer Ophthalmic Repository.

**Methods:** Randomly sampled clinical notes from glaucoma encounters dated 2014 to 2024 were labeled by two glaucoma specialists (with an adjudicator as needed) for prior glaucoma-related procedures (name, date, details) and recommended future procedures (name, details). Notes were split into development (10%), validation (10%), and test (80%) sets. Five LLMs (Claude Opus 4.6, DeepSeek-V4 Pro, GPT-5.5, Grok 4.1, and Qwen3.6-35B-A3B) were accessed in Microsoft Foundry through HIPAA-compliant containers. The extraction pipeline comprised a three-step agentic workflow (extraction, validation, and conditional revision) followed by text standardization. Detection of procedure data was evaluated using accuracy, sensitivity, specificity, precision, and F1-score. Among positive cases, performance was evaluated using exact-match accuracy and Jaccard Index (JI). Prior procedure labels were also compared with EHR glaucoma-related Current Procedural Terminology (CPT) codes.

**Main Outcome Measures:** Accuracy, exact-match accuracy, JI.

**Results:** A total of 1,984 (992 notes × 2 eyes) labels were evaluated for each task in the test set. Prior and future procedures were identified in 1,057 (53.3%) and 152 (7.7%) eyes, respectively. Accuracy ranged from 0.965 (Grok) to 0.996 (Claude) for prior procedures and 0.969 (DeepSeek, Qwen)-0.978 (Claude) for future procedures. Among positive cases, exact-match accuracy ranged from 0.681 (Grok) to 0.890 (Claude) for prior procedures and 0.704 (Grok)-0.849 (Claude) for future procedures; JI ranged from 0.827 (Grok) to 0.956 (Claude) and 0.734 (Grok) to 0.883 (Claude), respectively. Among all 1,760 prior procedures present in adjudicated labels, only 441 (25.1%) were captured by CPT-coded EHR data.

**Conclusions:** A secure agentic LLM pipeline accurately extracted glaucoma procedural data from clinical notes, recovering data largely absent from structured EHR fields. Claude performed best across both tasks. Limited agreement between labels and CPT codes likely reflects procedures performed at outside institutions and the limited granularity of CPT coding. This approach may support a scalable, comprehensive characterization of procedure burden among glaucoma patients for research and clinical applications.

---

Glaucoma is a chronic, progressive optic neuropathy and a leading cause of irreversible blindness worldwide, affecting tens of millions of individuals globally.^1^ Management typically extends across a patient’s lifetime and involves the use of medications, laser therapies, and incisional surgeries to slow disease progression.^2^ Procedural interventions not only aid in intraocular pressure (IOP) lowering, but may also blunt IOP fluctuations,^3,4^ which are theorized to influence disease progression.^5,6^ Two patients with an identical mean IOP may have meaningfully different disease trajectories depending on their procedural histories. For example, a clinician would likely assess the risk of disease progression differently for a patient with an IOP of 14 mmHg on four topical medications and a patient with the same IOP on no medications following trabeculectomy; the former depends greatly on adherence to a demanding medication regimen, whereas the latter may benefit from greater postoperative IOP stability.^7^ Because treatment burden accumulates and shifts over years, an accurate account of a patient’s treatment and procedural history is fundamental to clinical decision-making.

Capturing a truly comprehensive procedural history, spanning a patient’s entire course of care rather than a single institution’s records, is likely to become increasingly important in healthcare. As management shifts toward individualized, patient-centered care, clinicians will value this information to account for the wide variation in disease severity and treatment history among patients. Likewise, as artificial intelligence (AI) is integrated into clinical decision support, an accurate record of each patient’s course will be essential for both training and deploying these tools. Procedural history is also central to outcomes research, as studies evaluating surgical efficacy depend on accurate and sufficiently granular treatment data. However, obtaining a complete procedural history is challenging due to patient transitions between clinics and geographic relocation, which can lead to gaps in treatment documentation. This challenge is particularly accentuated at tertiary referral centers, where patients frequently present after extensive community-based treatment.

While electronic health records (EHRs) are a major source of clinical research data,^8^ procedural histories within a single EHR capture interventions performed only at that institution. Large multicenter resources, including the American Academy of Ophthalmology Intelligent Research in Sight (IRIS) Registry and the National Institutes of Health *All of Us* program, have expanded access to structured procedural data across diverse populations. However, such structured data rely on generic Current Procedural Terminology (CPT) codes, which may be incomplete and often lack the granularity needed for high-quality retrospective analyses. These repositories exclude free-text clinical notes, in part because unstructured data may contain protected health information (PHI).^9^ This exclusion is a substantial limitation: much clinically meaningful information is never captured in structured fields, with only approximately 13% of concepts documented in clinical notes having a comparable structured counterpart.^10^ In particular, procedures performed at outside institutions are only documented in narrative form. Clinical notes therefore represent a rich but largely underutilized source of procedural history. Although manual chart review can capture this information accurately, it is labor-intensive and difficult to scale.

Large language models (LLMs) offer a scalable approach to recover these data from free text. LLMs can interpret and extract content with high fidelity, expanding both the scope and reliability of automated extraction. Agentic workflows represent a further advance, structuring a task as a sequence of reasoning steps that incorporate conditional branching and self-correction. This approach has attracted considerable interest across clinical and research applications in medicine, particularly for complex tasks.^11^ Within ophthalmology, prior natural language processing (NLP) work on procedural data extraction has relied predominantly on rule-based approaches rather than LLMs.^12^ We have previously described how LLMs can be used in a HIPAA-compliant manner to extract risk factors, medication data, and classification of disease severity, type, and diagnosis from clinical notes in a standardized fashion.^13–15^

The purpose of this study was to evaluate the accuracy of LLMs in extracting standardized procedural information directly from free-text glaucoma clinical notes. In this study, we applied a three-step agentic workflow to extract procedural information followed by a text standardization step. We aimed to evaluate the accuracy of five different HIPAA-compliant LLMs in identifying each procedure, date, and associated clinical details for both procedural history and recommended future interventions. We hypothesized that a staged agentic workflow would achieve high accuracy in extracting both prior and recommended procedures, comparable to our previously reported performance in medication extraction. We also hypothesized that note-derived procedural data would be more complete than those derived from EHR CPT codes.

## Methods

### Data Collection

The University of Miami’s Institutional Review Board approved this study and waived the requirement for informed consent due to the retrospective nature of this work. This study followed the Declaration of Helsinki, and patient confidentiality was maintained in accordance with the Health Insurance Portability and Accountability Act (HIPAA).

This retrospective study analyzed patients from the Bascom Palmer Ophthalmic Repository (BPOR), a database encompassing more than 70,000 patients seen at the Bascom Palmer Eye Institute (BPEI). The methodology used for data extraction from the EHR system (Epic Systems, Verona, WI) has been described in our prior work.^16^ The database includes demographic, exam, imaging, clinical notes, and procedural records from BPEI clinical sites. Our methodology for procedural data extraction was similar to our prior published work.^15^ Briefly, clinical notes from outpatient encounters dated from 2014 to 2024 were extracted from BPOR. We only used notes fulfilling the following criteria: glaucoma-related encounters with notes signed by glaucoma specialist, glaucoma-related ICD code (H40.X) as a visit diagnosis, and patient age of 18 years or more. This yielded 451,283 eligible notes, from which we randomly sampled 1,250 notes corresponding to 1,250 distinct patients. For patients with multiple eligible encounters, a single note was selected at random. The resulting sample spanned 20 glaucoma specialists at BPEI. All notes were stored on a HIPAA-complaint, institutionally approved server (Box, Redwood City, CA). We further restricted the sample to clinically relevant visit types (e.g., new patient and follow-up visit encounters) and removed notes shorter than 200 characters to maintain data quality. This process revealed eight notes that were physician attestations rather than clinical documentation. These were excluded, leaving a final sample of 1,242 notes.

### Note Labeling

Clinical notes were independently labeled by two glaucoma specialists, with a third glaucoma specialist serving as an adjudicator. For each eye, graders extracted prior and recommended future procedures in three categories: glaucoma-related surgeries (trabeculectomy, aqueous shunt insertion, cyclodestructive procedures, microinvasive glaucoma surgeries), anterior segment lasers (iridotomy, iridoplasty, trabeculoplasty, YAG capsulotomy), and cataract surgery. Oculoplastic, cornea, and retina procedures were excluded. Prior procedures were labeled with the procedure name, date, and relevant procedural details such as the location of treatment, location of tube shunt quadrant or insertion site in the eye, degree of treatment, and number of treatments. Future recommended procedures were labeled with the procedure name and associated details. If laterality of the procedure was unclear, labelers were instructed to add the phrase “(laterality unknown)” to the labels and assign the procedure to both eyes.

### CPT-derived Procedural History Extraction

CPT-encoded procedural history was extracted from the EHR for each of the 1,242 subjects. CPT codes corresponding to the procedures noted above were extracted. We excluded any procedures occurring after the encounter date of the associated clinical note of each subject. CPT codes were mapped to standardized procedure names (see below for further details on text standardization). Procedure laterality was determined using the associated laterality modifier in the EHR database. When this field was not documented, laterality was inferred from the associated ICD-10 code.

### LLM Prompt Engineering and Agentic Workflow

The dataset was split into three sets stratified by clinician: development (10%), validation (10%), and held-out test set (80%).^15^ The development set was used to guide prompt engineering and provide few-shot examples, while the validation set was used to refine prompt structure and rules prior to final evaluation. The LLM was not fine-tuned, allowing for a relatively small development set compared to traditional supervised learning as previously discussed.^15^

Separate LLM extraction pipelines were created for prior and future procedures. Each consisted of a sequential three-stage agentic workflow: initial extraction, validation, and conditional revision (Figure 1). Prompts incorporated rule-based instructions, hierarchical evidence prioritization, few-shot examples, checklist-guided self-verification, and structured reasoning decomposition inspired by chain-of-thought prompting. The clinical note and encounter date served as input and models were instructed to return a structured JSON output. The initial extraction step generated the procedure label along with supporting citations and reasoning. This output and the original clinical note were then passed to the validation step, which either confirmed correct extraction or flagged a disagreement. If validation passed, the initial output was retained. If not, the note and initial output were passed to the conditional revision step, which was instructed to revise the output using the same prompt provided in the initial extraction step. The full text of prompts is available in the Supplemental Material (available at https://www.ophthalmologyscience.org).

**Figure 1.**
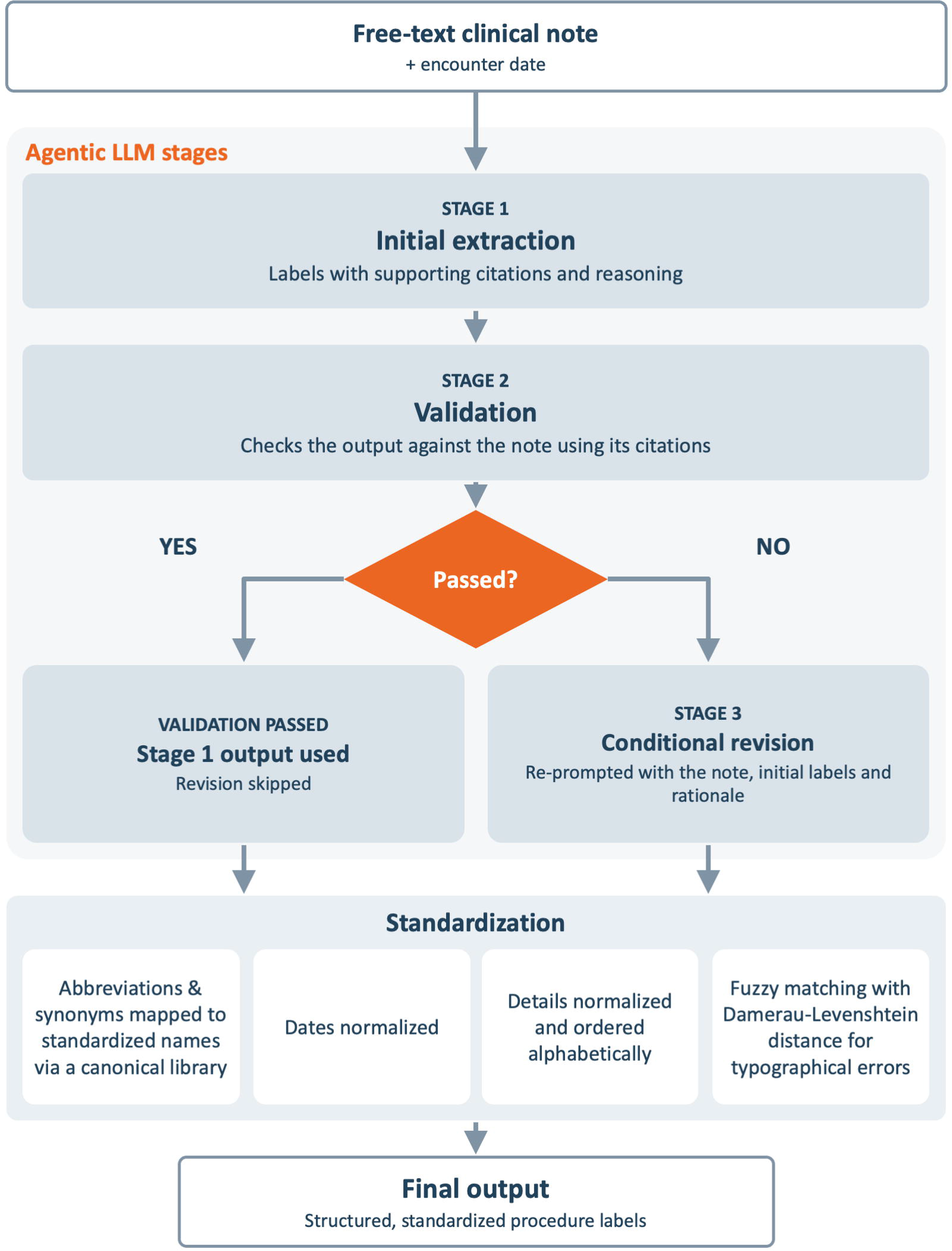
Workflow of the agentic large language model and standardization pipeline.

Detailed explanation of LLM failure handling is described in our prior work.^15^ Briefly, three failure types were defined: 1) model failures, in which outputs were truncated at the maximum token limit with no label content recoverable through a tiered JSON and regular expression (regex) parsing strategy; 2) API timeout failures, in which a model did not respond within the allotted time window; and 3) standardization failures, in which an extracted procedure name could not be mapped to the canonical procedure dictionary, resulting in fallback to the input label. All failures were retained in performance metrics.

### LLM Access

LLMs were accessed through a secure cloud-based environment using Microsoft Foundry (Microsoft, Redmond, WA). An established business associate agreement between Microsoft and the University of Miami allowed access to HIPAA-compliant containers where all LLM tasks were run. We chose to run our pipeline on five leading LLMs: Claude Opus 4.6, DeepSeek-V4 Pro, GPT-5.5, Grok 4.1 fast non-reasoning, and Qwen3.6-35B-A3B. We ran all model experiments with the same parameters: temperature of 0.4, top_p of 0.9, and maximum token length of 750. The reasoning effort was set to “none” for all models to ensure a fair comparison. “None” was available for all models except DeepSeek, which used “minimal” as its lowest reasoning setting.

### Label and Output Standardization

To enable the direct comparison between adjudicated labels, LLM-generated outputs, and CPT-encoded procedural data, we built a Python script to standardize all text prior to performance evaluation. Each output was parsed into three components: procedure name, procedural details (in parentheses, if available), and date (if available), with any extraneous text removed. Procedure names were mapped using a canonical dictionary that accounted for common abbreviations and synonyms. Clinically relevant procedural details were retained in parenthesis and listed alphabetically. Dates were standardized only for prior procedures since future procedures were not associated with a specific date. When there was uncertainty about whether a procedure occurred or about its identity, “possible” was appended as a prefix to the procedure name. When the date of a prior procedure was uncertain, a tilde (“∼”) was appended to the date to indicate approximation. Typographical errors in procedure name were corrected using a fuzzy matching algorithm based on the Damerau-Levenshtein distance, which quantifies the number of insertions, deletions, substitutions, and transpositions required to convert one string into another.^17,18^ If multiple procedures were listed, they were ordered first chronologically, then alphabetically. The code for the LLM pipeline and standardization program, including a Dockerfile for containerized execution, is available on GitHub: https://github.com/swarupswaminathan/llm_procedures.

### Comparison with Structured EHR Procedural History

To compare note-derived procedural data with structured EHR records, CPT-encoded procedural history was compared with adjudicated labels. Because CPT codes are designed for billing, they are less specific than procedural descriptions in clinical notes. A single code may encompass multiple procedures or device types and lack details such as the specific implant used. Direct comparison with adjudicated labels may therefore underestimate agreement when both sources describe the same procedure at different levels of granularity. To address this asymmetry, we mapped CPT codes to procedure categories. Note-derived procedure labels were then mapped to this broader category level to evaluate agreement with CPT-logged procedures. For example, CPT code 66180 recorded on 12/1/2015 would map to the procedure category of “tube shunt.” An adjudicated label documenting “Ahmed FP7 12/1/2015” would be assigned to the same procedure category and considered a match. Within each eye, adjudicated label and CPT-coded procedures were first matched by procedure category. When more than one candidate matched, procedures were matched by the closest date. Each procedure was then classified as present in both data sources, note only, or CPT-code only for further analysis.

### Statistical Analysis

We used Gwet AC1 to assess intergrader agreement for both tasks. For this analysis, we initially treated labels as binary, with an empty label considered “No”, and a label containing one or more procedures considered “Yes” (e.g., “trabeculectomy 3/6/2019” or “possible SLT”). Similarly, to evaluate model performance, we first assessed agreement between the standardized adjudicated labels and LLM outputs regarding the presence or absence of procedure data using Gwet AC1 as well as specificity, sensitivity (recall), precision, and F1 score.

Among positive cases, we then evaluated text-level accuracy using exact match accuracy and the Jaccard Index (JI). JI quantifies overlap between two sets as the size of their intersection divided by the size of their union and is commonly used in data science to compare predicted and reference classifications.^19,20^ Values range from 0 (no overlap) to 1 (complete overlap). We compared full procedure entries rather than individual words to avoid inflating agreement from partial or scrambled matches. Partial matching on dates was allowed, but procedure names had to match fully before credit for partial dates was awarded. For each matched procedure entry, we calculated overlap across procedure name, details, and decomposed date components (day, month, year). Procedure name and detail components were weighted as 33% each, while individual date components were each weighted as 11%, such that a fully correct date contributed the same total weight as the name or detail component. For example, if the adjudicated label was “BGI (sulcus) 5/5/2020” and the LLM output was “BGI (sulcus) 5/2020”, the calculation will credit the matching name, details, month and year, while penalizing for the missing day information, resulting in a JI value of 0.89. Within the positive cases, exact match accuracy and JI were also computed for subsets of each label – for the prior procedure task, procedure name and date (excluding details) and procedure name alone; for the future procedure task, procedure name alone (excluding details).

95% confidence intervals (CIs) for F1 score and JI were generated using 1,000 bootstrap resamples, while CIs for specificity, sensitivity, and precision were calculated using the Wilson method. Failure rates were tabulated for each model and task. Failed outputs resulted in a fallback of “None” and were retained in all performance metrics. Finally, labels and LLM outputs for both tasks were converted into a count of procedures per note and mean absolute error (MAE) was calculated between the LLM-generated and adjudicated counts. To account for potential bias from notes without documented procedures, MAE was computed both across all notes and among positive cases only.

For comparison between note-derived labels and CPT-based procedural data, agreement was summarized overall and by procedure type using three measures: the proportion of CPT-coded procedures documented in the notes, the proportion of note-derived procedures captured by CPT-coded logs, and the positive agreement. Positive agreement was defined as the proportion of total positive procedure records across both sources that were concordant, calculated as 2*a* / (2*a* + *b* + *c*). Here, *a* represents procedures identified in both clinical notes and CPT records, while *b* and *c* represent procedures identified exclusively in the notes or exclusively in the CPT records, respectively. Positive agreement is mathematically equivalent to the F1 score and is symmetric with respect to which source is treated as the reference. We also measured date agreement at four levels: exact match, within 7 days, same month, and same year. We took this additional step to account for potential typographical errors with date entry as well as clinicians deliberately excluding dates in their notes due to perceived minimal clinical relevance.

## Results

A total of 1,984 labels (at the eye level; two labels per note) were evaluated in the test set for prior and future procedure data. Gwet AC1 for intergrader agreement was 0.875 for prior and 0.964 for future labels. Adjudications were required for 673 (33.9%) prior procedure labels and 91 (4.6%) future procedure labels. A total of 1,057 (53.3%) eyes had procedural history, and 152 (7.7%) eyes received recommendations for future procedures. The median number of prior procedures in an eye was 1 (IQR 0-1), with a mean of 0.9±1.1. The median number of future procedures in an eye was 0 (IQR 0-0), with a mean of 0.1±0.4 (Figure 2).

**Figure 2.**
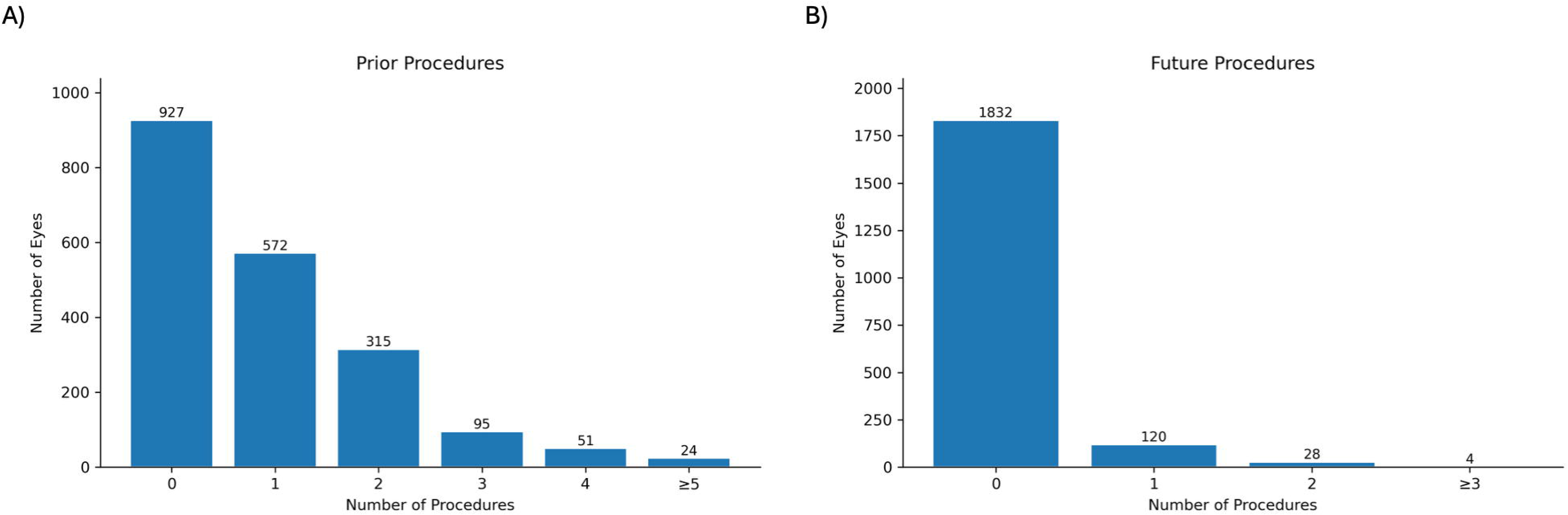
Distribution of procedure counts per eye for (A) prior procedures and (B) future procedures per reference labels.

Table 1 displays the LLM failure rates and performance in identifying the presence or absence of procedural data pertinent to each task. All models experienced low failure rates, averaging 1.9% for prior procedures and 0.3% for future procedures. Most failures were standardization failures; no model failures occurred. Stage 3 was executed at an average of 173 times (17.5% of notes) across all models for prior procedures, and an average of 56 times (5.6% of notes) for future procedures. With the inclusion of Stages 2 and 3 in the workflow, exact match accuracy in positive cases improved by an average of 7.7% (ranging from 1.9% for GPT to 14.9% for DeepSeek) in prior tasks and 1.9% (ranging from-2.8% for Grok to 5.7% for Claude) in future tasks. The standardization script successfully fuzzy matched procedure names an average of 7.0 times (0.2%) in prior tasks and an average of 1.2 times (0.3%) in future tasks.

**Table 1.** Overall model performance metrics utilizing different large language models for procedure-related tasks.

|  |  | Claude | DeepSeek | GPT | Grok | Qwen |
| --- | --- | --- | --- | --- | --- | --- |
| **Prior** | Failure rate | 1.46% (S-29) | 3.02% (A-2; S-58) | 1.56% (S-31) | 1.46% (A-2; S-27) | 1.92% (S-38) |
|  | Accuracy | 0.996 (0.993 - 0.998) | 0.982 (0.975 - 0.987) | 0.992 (0.988 - 0.995) | 0.965 (0.956 - 0.972) | 0.977 (0.970 - 0.983) |
|  | Gwet AC1 | 0.993 (0.987 - 0.998) | 0.964 (0.953 - 0.975) | 0.985 (0.977 - 0.992) | 0.930 (0.914 - 0.945) | 0.955 (0.943 - 0.968) |
|  | Specificity | 0.999 (0.994 - 1.000) | 0.989 (0.980 - 0.994) | 0.999 (0.994 - 1.000) | 0.997 (0.991 - 0.999) | 0.989 (0.980 - 0.994) |
|  | Sensitivity (recall) | 0.994 (0.988 - 0.997) | 0.975 (0.964 - 0.983) | 0.987 (0.978 - 0.992) | 0.937 (0.920 - 0.950) | 0.967 (0.954 - 0.976) |
|  | Precision | 0.999 (0.995 - 1.000) | 0.990 (0.982 - 0.995) | 0.999 (0.995 - 1.000) | 0.997 (0.991 - 0.999) | 0.990 (0.982 - 0.995) |
|  | F1 score | 0.997 (0.994 - 0.999) | 0.983 (0.977 - 0.988) | 0.993 (0.989 - 0.996) | 0.966 (0.958 - 0.974) | 0.978 (0.972 - 0.984) |
|  | MAE | 0.024 (0.017 - 0.031) | 0.092 (0.079 - 0.106) | 0.045 (0.035 - 0.055) | 0.142 (0.124 - 0.161) | 0.099 (0.084 - 0.114) |
| **Future** | Failure rate | 0.20% (S-4) | 0.45% (A-2; S-7) | 0.40% (A-1; S-7) | 0.25% (A-1; S-4) | 0.10% (S-2) |
|  | Accuracy | 0.978 (0.971 - 0.984) | 0.969 (0.961 - 0.976) | 0.975 (0.967 - 0.981) | 0.975 (0.967 - 0.981) | 0.969 (0.960 - 0.976) |
|  | Gwet AC1 | 0.974 (0.966 - 0.982) | 0.964 (0.954 - 0.973) | 0.971 (0.962 - 0.979) | 0.970 (0.962 - 0.978) | 0.963 (0.954 - 0.973) |
|  | Specificity | 0.981 (0.974 - 0.987) | 0.974 (0.965 - 0.980) | 0.980 (0.972 - 0.985) | 0.988 (0.982 - 0.992) | 0.978 (0.970 - 0.983) |
|  | Sensitivity (recall) | 0.941 (0.891 - 0.969) | 0.914 (0.859 - 0.949) | 0.921 (0.867 - 0.954) | 0.816 (0.747 - 0.869) | 0.862 (0.798 - 0.908) |
|  | Precision | 0.808 (0.744 - 0.859) | 0.743 (0.676 - 0.801) | 0.791 (0.725 - 0.844) | 0.849 (0.782 - 0.898) | 0.762 (0.693 - 0.819) |
|  | F1 score | 0.869 (0.830 - 0.905) | 0.820 (0.775 - 0.861) | 0.851 (0.807 - 0.891) | 0.831 (0.785 - 0.875) | 0.808 (0.760 - 0.850) |
|  | MAE | 0.034 (0.023 - 0.045) | 0.042 (0.031 - 0.053) | 0.033 (0.023 - 0.044) | 0.034 (0.024 - 0.043) | 0.043 (0.032 - 0.054) |

MAE = mean absolute error.
Failure-rate notation: A = application programming interface (API) timeout failure and S = standardization failure, with the number indicating the number of labels that failed for each etiology.

Accuracy of binary assessment for the prior procedure task ranged from 0.965 (Grok) to 0.996 (Claude). F1 scores ranged from 0.966 (Grok) to 0.997 (Claude). Claude and GPT performed the best across all metrics in the prior procedure task. Accuracy for the future procedure task ranged from 0.969 (DeepSeek, Qwen) to 0.978 (Claude). F1 scores ranged from 0.808 (Qwen) to 0.869 (Claude). Distribution of errors in the number of procedures for both tasks is displayed in Figure 3.

**Figure 3.**
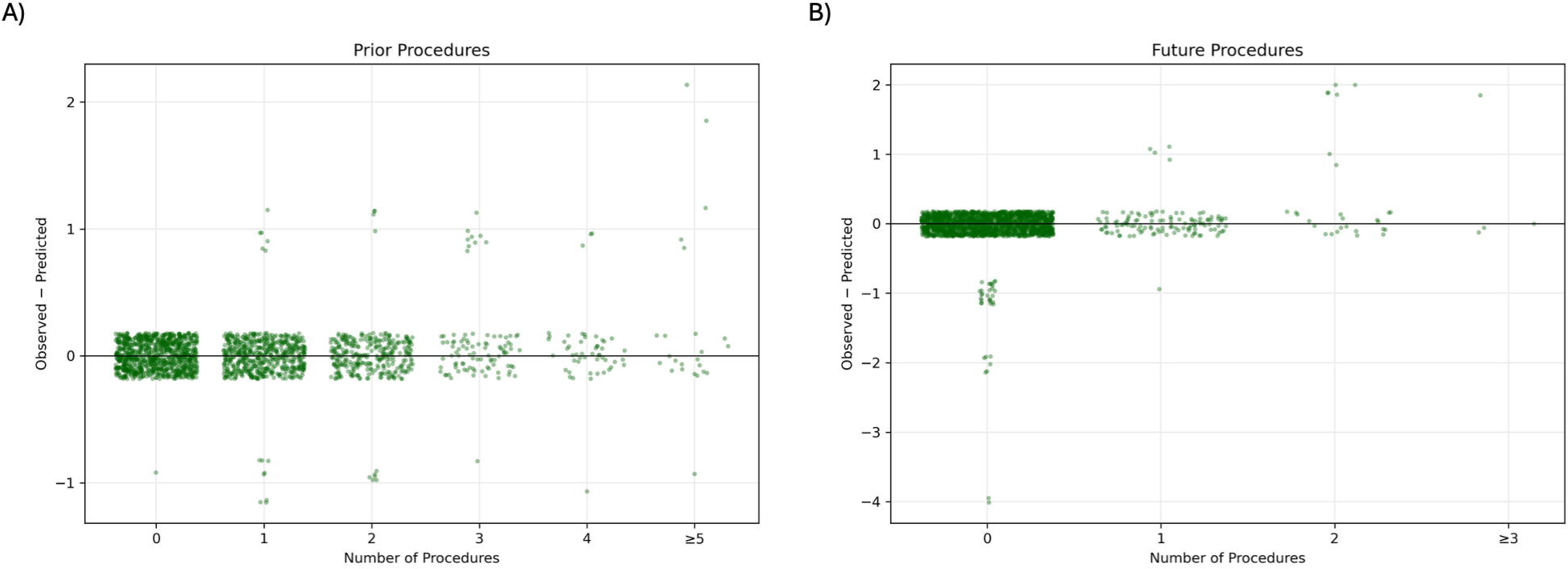
Sina plots of procedure count prediction error for (A) prior procedures and (B) future procedures using Claude outputs. Points are jittered horizontally in proportion to local point density and vertically for visibility.

Table 2 displays text-level analysis among positive cases. Exact match accuracy for the prior procedure task ranged from 0.681 (Grok) to 0.890 (Claude). JI ranged from 0.827 (Grok) to 0.956 (Claude). Among positive cases, MAE for number of prior procedures in the output ranged from 0.043 (Claude) to 0.265 (Grok). Exact match accuracy for the future procedure task ranged from 0.704 (Grok) to 0.849 (Claude). JI ranged from 0.734 (Grok) to 0.883 (Claude). Positive-case MAE for number of future procedures in the output ranged from 0.105 (GPT) to 0.263 (Grok). Claude achieved the highest JI and exact match accuracy for both tasks; GPT ranked second for prior procedures and had the lowest MAE for future procedures. Component-level performance among positive cases is shown in Table 3. For the prior procedure task, when procedure name and date (excluding details) were evaluated, exact match accuracy ranged from 0.710 (Grok) to 0.917 (Claude). JI ranged from 0.834 (Grok) to 0.961 (Claude). Model performance was also computed for procedure name only in the prior procedure task. Exact match accuracy ranged from 0.755 (Grok) to 0.942 (Claude), and JI ranged from 0.852 (Grok) to 0.970 (Claude). For the future procedure task, model performance was measured for procedure name only (excluding details). Exact match accuracy ranged from 0.743 (Grok) to 0.901 (Claude), and JI ranged from 0.757 (Grok) to 0.911 (Claude). Across all component-level analyses, Claude achieved the best performance.

**Table 2.** Model performance metrics among positive cases only for each task.

|  |  | Claude | DeepSeek | GPT | Grok | Qwen |
| --- | --- | --- | --- | --- | --- | --- |
| **Prior** | Jaccard Index | 0.956 (0.947 - 0.965) | 0.881 (0.866 - 0.896) | 0.918 (0.905 - 0.930) | 0.827 (0.809 - 0.845) | 0.851 (0.835 - 0.867) |
| **N=1057** | Exact match accuracy | 0.890 (0.870 - 0.908) | 0.737 (0.710 - 0.763) | 0.815 (0.790 - 0.837) | 0.681 (0.652 - 0.709) | 0.692 (0.663 - 0.719) |
|  | MAE | 0.043 (0.031 - 0.057) | 0.161 (0.137 - 0.187) | 0.084 (0.068 - 0.102) | 0.265 (0.233 - 0.298) | 0.177 (0.149 - 0.202) |
| **Future** | Jaccard Index | 0.883 (0.832 - 0.928) | 0.845 (0.790 - 0.898) | 0.842 (0.786 - 0.893) | 0.734 (0.665 - 0.800) | 0.774 (0.710 - 0.834) |
| **N=152** | Exact match accuracy | 0.849 (0.783 - 0.897) | 0.809 (0.739 - 0.864) | 0.816 (0.747 - 0.869) | 0.704 (0.627 - 0.771) | 0.750 (0.676 - 0.812) |
|  | MAE | 0.125 (0.059 - 0.197) | 0.135 (0.066 - 0.211) | 0.105 (0.053 - 0.158) | 0.263 (0.178 - 0.355) | 0.217 (0.145 - 0.303) |

MAE = mean absolute error.

**Table 3.**
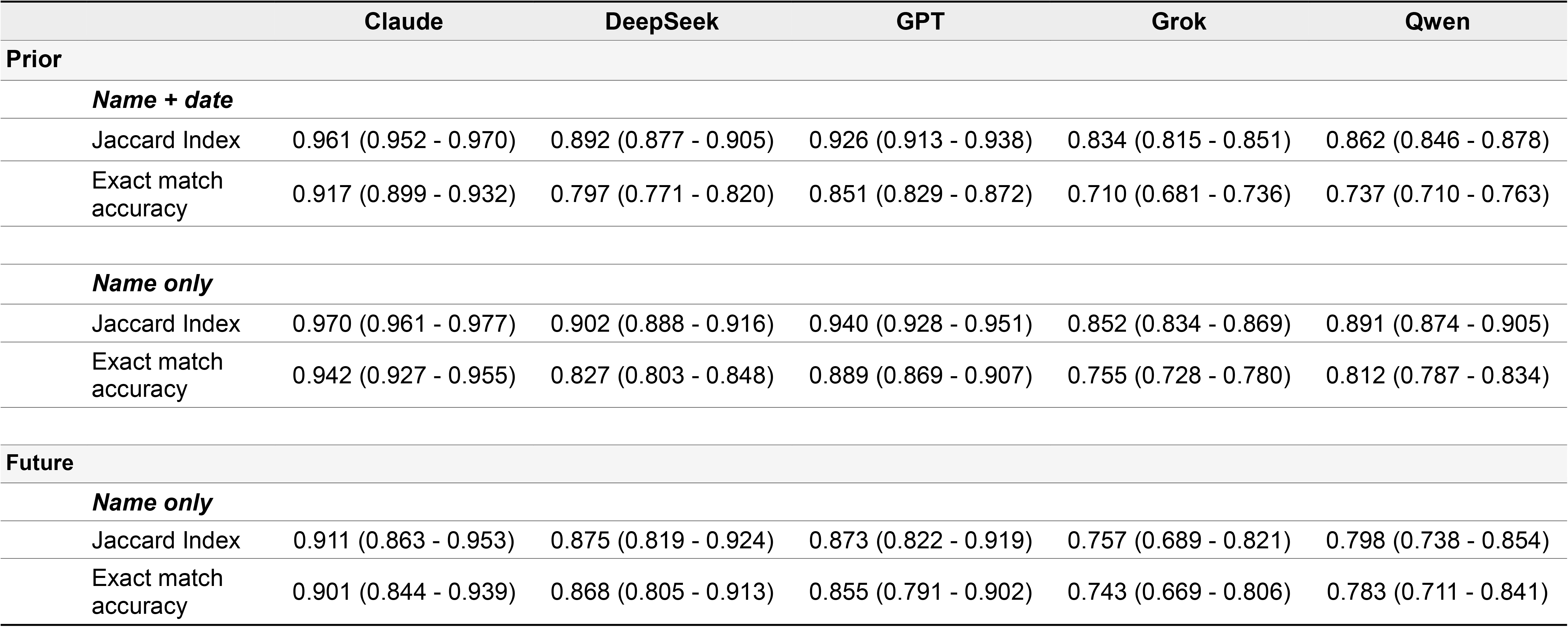
Model performance metrics for subsets of label components among positive cases.

|  | Claude | DeepSeek | GPT | Grok | Qwen |
| --- | --- | --- | --- | --- | --- |
| Prior |  |  |  |  |  |
| Name + date |  |  |  |  |  |
| Jaccard Index | 0.961 (0.952 - 0.970) | 0.892 (0.877 - 0.905) | 0.926 (0.913 - 0.938) | 0.834 (0.815 - 0.851) | 0.862 (0.846 - 0.878) |
| Exact match accuracy | 0.917 (0.899 - 0.932) | 0.797 (0.771 - 0.820) | 0.851 (0.829 - 0.872) | 0.710 (0.681 - 0.736) | 0.737 (0.710 - 0.763) |
| Name only |  |  |  |  |  |
| Jaccard Index | 0.970 (0.961 - 0.977) | 0.902 (0.888 - 0.916) | 0.940 (0.928 - 0.951) | 0.852 (0.834 - 0.869) | 0.891 (0.874 - 0.905) |
| Exact match accuracy | 0.942 (0.927 - 0.955) | 0.827 (0.803 - 0.848) | 0.889 (0.869 - 0.907) | 0.755 (0.728 - 0.780) | 0.812 (0.787 - 0.834) |
| Future |  |  |  |  |  |
| Name only |  |  |  |  |  |
| Jaccard Index | 0.911 (0.863 - 0.953) | 0.875 (0.819 - 0.924) | 0.873 (0.822 - 0.919) | 0.757 (0.689 - 0.821) | 0.798 (0.738 - 0.854) |
| Exact match accuracy | 0.901 (0.844 - 0.939) | 0.868 (0.805 - 0.913) | 0.855 (0.791 - 0.902) | 0.743 (0.669 - 0.806) | 0.783 (0.711 - 0.841) |

Adjudicated labels in the test set contained a total of 1,760 standardized prior procedures, while EHR-derived procedural data contained 548 CPT-encoded procedures. Figure 4 depicts the matching approach used and frequency of various CPT codes in this cohort. A total of 441 procedures were found in both sources. Of the CPT-coded procedures, 80.5% were documented in notes, whereas only 25.1% of the note-derived procedures were captured by CPT data for an overall positive agreement of 38.2% (Table 4). Among the 441 matches, dates from adjudicated labels and CPT procedural data agreed exactly in 289 cases (65.5%), within 7 days in 301 cases (68.3%), within the same month in 336 cases (76.2%), and within the same year in 363 cases (82.3%). The remaining note-derived procedures had no associated date. Of the 107 CPT-listed procedures that did not categorically match a labeled procedure (i.e., not documented in the clinic note), most common were capsulotomy (32.7%), iridotomy (16.8%), trabeculoplasty (12.1%), and cataract surgery (11.2%). Remaining cases mostly involved discrepancies in tube shunt versus tube revision procedures.

**Figure 4.**
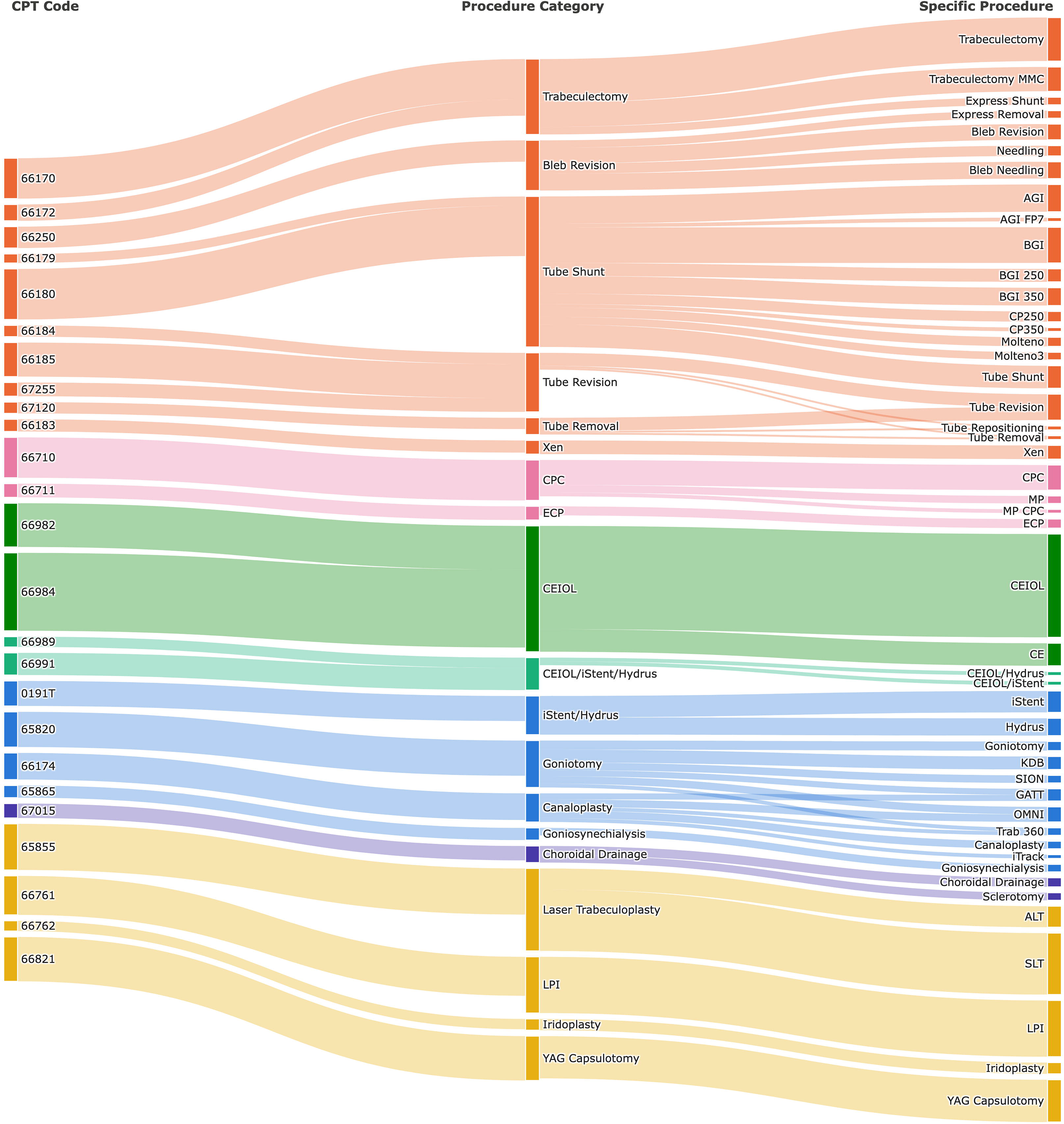
Sankey diagram depicting the mapping of CPT-encoded procedures with note-derived procedure labels. Thickness of bands reflect frequency in the data set, scaled as 1+√n to maintain the visibility of infrequent codes.

**Table 4.** Agreement between note-derived and Current Procedural Terminology (CPT)-coded procedures, overall and by procedure category.

| Procedure Type | Note-derived, n | CPT-coded, n | In both, n | CPT procedures documented in note, % | Note procedures captured by CPT codes, % | Positive agreement, % |
| --- | --- | --- | --- | --- | --- | --- |
| *Overall* | *1760* | *548* | *441* | *80.5* | *25.1* | *38.2* |
| Cataract surgeries | 727 | 156 | 144 | 92.3 | 19.8 | 32.6 |
| Lasers | 561 | 210 | 144 | 68.6 | 25.7 | 37.4 |
| Filtering conjunctival surgeries | 373 | 120 | 100 | 83.3 | 26.8 | 40.6 |
| Microinvasive glaucoma surgeries | 61 | 40 | 35 | 87.5 | 57.4 | 69.3 |
| Cyclodestructive procedures | 35 | 18 | 16 | 88.9 | 45.7 | 60.4 |
| Choroidal drainage | 3 | 4 | 2 | 50.0 | 66.7 | 57.1 |

## Discussion

In this study, an agentic LLM pipeline accurately extracted both prior and recommended future glaucoma procedures from free-text clinical notes when compared with an expert-labeled reference standard. While extracting procedural data from unstructured glaucoma clinical notes has traditionally required labor-intensive manual chart review, our study demonstrates that an LLM-based pipeline can accomplish this task both accurately and at scale. Importantly, much of the procedural history recovered by the pipeline was absent from structured EHR data. A standardization step was critical to this performance given the heterogeneity of free-text documentation. Mapping procedure names to a canonical dictionary of abbreviations and synonyms, correcting typographical errors with fuzzy matching, and normalizing dates converted variable notation into consistent, analyzable outputs. Performance across models was strong, with several LLMs reaching high exact match accuracy and JI values against the reference standard. Claude achieved the highest accuracy, F1 score, JI, and exact match accuracy for both tasks. All processing was performed within a HIPAA-compliant environment using Microsoft Foundry, supporting the feasibility of deploying this approach within existing institutional frameworks. These findings suggest that agentic LLM pipelines can convert unstructured clinical notes into structured procedural data at a scale not feasible with manual review, helping address a persistent challenge in procuring all available data for clinical care, outcomes research, and AI algorithm development.

Model performance varied by task, with greater performance for prior than future procedure extraction for most models. This difference was likely due to the smaller number of positive cases in the future procedure task. More than half of eyes in the study cohort had undergone a prior procedure, while less than 10% had a documented proposed future procedure. For prior procedures, exact match accuracy and JI improved across all models when parenthetical details were excluded, with JI increasing to 0.961 with Claude, for example. This pattern demonstrates the strong ability of our LLM pipeline to correctly identify the type of procedure with its associated date, which are arguably the most clinically significant components. Procedural details, such as the quadrant of a tube shunt or the degrees of angle treated with SLT, are useful but are less essential to characterizing a patient’s overall clinical history. For most clinical and research applications such as modeling disease progression, knowing which procedures a patient has undergone and when they were performed provide the most valuable information. Errors confined to procedural details are therefore less likely to meaningfully affect downstream analyses.

Supplemental Table 5 (available at https://www.ophthalmologyscience.org) demonstrates examples of LLM failure for both tasks. Failures in prior procedure extraction arose mainly from ambiguity within the note itself, date discrepancies, and differing interpretations of clinical uncertainty. Many date extraction failures followed this pattern of multiple dates in the note with varying degrees of specificity and occasional conflicting dates likely attributable to typographical errors rather than true model errors. For the future procedure task, failures were most often due to difficulty distinguishing definitive clinical recommendations from tentative considerations or conditional suggestions. In most examples, the conditional revision step (Stage 3) was triggered, with LLM reasoning indicating that the model recognized ambiguity in the strength of the recommendation.

A key long-term goal in glaucoma care is to leverage advances in data science and AI to individualize management according to each patient’s disease characteristics. AI models for glaucoma progression will certainly draw on known risk factors such as age, race, family history, intraocular pressure, as well as visual field and optical coherence tomography data. However, treatment burden is rarely captured with the same precision, even though it will likely influence clinical trajectory and prognosis as observed in ophthalmology and other fields.^21–23^ Accurate knowledge of prior procedural history is essential for guiding clinicians and patients in considering treatment options. Building on our prior work extracting medication data and risk factors such as family history and adherence,^14,15^ this pipeline adds procedural history to create a comprehensive, structured representation of risk factors and treatment burden extracted from clinical notes, complementing existing structured EHR data. This information could improve patient characterization in the training and deployment of AI models, potentially enhancing predictive endpoints such as the probability of glaucoma progression.^24^ Extending this LLM-based extraction pipeline to additional institutions could enrich the procedural data available in large databases such as the IRIS Registry. The pipeline could be deployed locally to extract and share de-identified output alongside existing structured data, thereby including procedures performed at external practices. These additional data would substantially strengthen the quality and scope of research that these databases can support.

One of the main sources currently used to obtain procedural history is the EHR CPT-derived procedural log. In our cohort, CPT data captured less than a quarter of procedures documented in clinical notes, with an overall positive agreement of 38.2%. This low capture persisted even after we mapped both sources to generic procedure categories, indicating that the gap reflects procedures absent from the structured data rather than differences in granularity alone. Several factors likely contribute to this finding. As noted, over the course of their chronic disease, patients may relocate or move between clinics, including transfers of care to tertiary care centers, and procedures performed at outside institutions are not captured in the primary institution’s billing data. In addition, procedures completed at the primary institution prior to the introduction of the EHR would be absent. Aside from challenges with completeness, CPT codes lack the granularity that clinical notes can provide. CPT codes map to procedures generically, such as “tube shunt” or “goniotomy,” even though each can be performed with numerous devices that differ in surgical success. Because large-scale retrospective studies of surgical outcomes often require identifying the use of a specific device, CPT-derived data alone are insufficient, whereas LLM-based extraction from clinical notes can capture this level of detail. For example, an LLM can extract output can capture specific procedure information such as “Baerveldt 350” instead of “tube shunt,” or “GATT” instead of “goniotomy”. A researcher comparing the effectiveness of different tube shunts, for instance, could first use CPT codes to identify patients who received a tube shunt at the institution and then apply an LLM pipeline to determine the specific device used in each case for further analysis.

Nonetheless, EHR-derived CPT procedural history has a key strength. Because CPT codes are used for billing, a coded procedure almost certainly occurred on the recorded date. In contrast, clinical notes omitted 19.5% of CPT-coded procedures, most of which were laser procedures. YAG capsulotomies and cataract surgeries may have been omitted from the note because the clinician did not consider them to be directly relevant to their patient’s glaucoma. Iridotomies may lose clinical relevance after cataract extraction, and trabeculoplasties after their effect wears off, so both may be documented less consistently over time. These procedures are also characterized by minimal anatomic findings. Note-derived dates were also less precise, with progressively greater agreement when matched within 7 days, by month, and by year. This pattern suggests that clinicians often record approximate dates, consistent with the general timing of a procedure mattering more clinically than its exact date. Ultimately, EHR CPT-derived data and notes may have complementary strengths – reliable occurrence and dates from CPT and procedural detail and external procedures from clinical notes. Combining them may provide the most complete picture of procedural history.

Prior work in NLP-based data extraction from ophthalmology clinical notes has largely relied on rule-based algorithms rather than LLMs^12,25^, or has focused on components of the clinical picture other than procedural data, such a medications,^15^ risk factors,^14^ or diagnoses.^13,26^ LLM-based extraction of procedural information specifically has begun to gain traction in other areas of medicine, such as orthopedic surgery,^27^ but has not been previously applied to glaucoma or broader ophthalmology research likely due to challenges in text standardization.

We acknowledge limitations to our methodology. This pipeline relies on clinical notes from glaucoma specialists and adult patients, with one clinical note evaluated per patient. While a single note may not capture information documented elsewhere in a patient’s record, procedural data is relatively static relative to medication regimens, making longitudinal data less critical for this task than for medication extraction. Because this study was conducted at a single institution, procedural abbreviations in the clinical notes may reflect institution-specific conventions that could limit generalizability to other settings. However, these differences could be accommodated by expanding the canonical procedure abbreviation library and including few-shot examples in the LLM prompts to reflect local documentation practices. Additionally, our pipeline requires access to Microsoft Foundry, a resource currently more accessible to larger academic institutions, though we expect this barrier to ease as such infrastructure becomes more widely available. While a standardization step reduced variability in extracted outputs, final performance remained dependent on the accuracy of both model extraction and post-processing, and we did not evaluate the effect of varying reasoning levels within each LLM given our focus on direct model comparison.

Future work includes validation of this pipeline at other institutions. This approach could also be extended to non-glaucoma encounters, pediatric patients, or other sub-specialties within ophthalmology. We could also pair LLM-extracted procedural data with CPT-derived procedural history, as mentioned above, to construct a more complete longitudinal procedural history for each patient. Given that our group has separately developed a similar pipeline for medication extraction,^15^ combining both into a single unified workflow represents a promising direction, potentially enabling simultaneous extraction of procedural and medication information.

In conclusion, this study demonstrates that an LLM-based agentic workflow can accurately and efficiently extract both prior and recommended procedural data from unstructured glaucoma clinical notes, achieving strong agreement with expert-labeled reference standards while substantially outperforming existing CPT-based procedural EHR data. By transforming free-text documentation into structured, standardized procedural data, this approach addresses a longstanding gap in the granularity and completeness of procedural history available for both clinical care and research. With validation at other institutions and use in other patient populations, this LLM-based workflow holds significant potential to improve the scale and precision with which procedural data can be extracted and used to support more individualized, data-driven glaucoma care.

## Declaration of Generative AI and AI-assisted technologies in the writing process

During the preparation of this work the authors used Claude to improve structural organization of manuscript text. After using this tool/service, the authors reviewed and edited the content as needed and take full responsibility for the content of the publication.

## Supporting information

Supplemental Material: Examples

Supplemental Material: Prompts

## Abbreviations

API: Application Programming Interface
EHR: Electronic Health Record
LLM: Large Language Model
NLP: Natural language processing
PHI: Protected Health Information

## Data Availability

All data produced in the present study are available upon reasonable request to the authors

https://github.com/swarupswaminathan/llm_procedures

