## Supplemental Material: Examples for "Beyond Billing Codes: An Agentic Large Language Model Pipeline for Extracting Glaucoma Procedural Data from Free-Text Clinical Notes"

**Table 5.** Examples of failures in LLM outputs for both procedure tasks. Pertinent note text is highlighted in red.

| Task | Note Text | Adjudicated label OD | Adjudicated label OS | LLM output OD | LLM output OS | Claude LLM reasoning |
|---|---|---|---|---|---|---|
| Prior | Impression: 1. Primary open angle glaucoma, original dx ~****. S/p SLT 360 OS in 2010 (Dr. ***). Elevated IOP noted **** and followed without treatment until ****. Untreated IOP by applanation ranged 25-29. On topical prostaglandin, IOP usually mid to upper teens. Meds advanced in ****. Dr. *** did 360 SLT left eye in ****. Resumed monotherapy in ****, with IOP usually upper teens. *** **** had disc hemorrhage OS and brimonidine OS was added to Travatan OU; she was intolerant and Azopt OS was substituted. She used Cosopt in the past with no comment regarding tolerability. Diode laser done OS nasally **.2016. She was sent back in **** to consider laser OD. There is a visit note on **.**, but that visit was signed out as patient left because she never saw the doctor. Her note had been set up and should be disregarded. Dr. *** records are in the Media tab as External health record, Visual fields/photos Today, **/**/****: ¬∑ IOP OD 20 / IOP OS 12 at **:** AM Applanation Using timolol 1/1 HVF OD mildly depressed compared to ****; OS is better than **** Undilated exam, history, and fields were sufficient for me to agree with Dr. *** regarding trabeculooplasty OD, and we chose to go ahead today and do dilation on visit to assess effect of laser Nasal diode trabeculoplasty done OD without problem 2. Nuclear cataracts OU 3. ?Scleroderma Dr. *** requested that we arrange a rheumatology evaluation. She went to see rheumatologist on **/**/**** with a diagnosis of limited scleroderma. They recommended follow-up in 2-3 months, but she didn't go back due to insurance issues. ............................................................................. Disposition Return in about 6 weeks (around **/**/****) for FUV slot, IOP OU with proparacaine HCl 0.5% one drop OU, dilate tropicamide1%, 1 drop each eye. Add steroid 4,4,4,2,2,2 to OD ¬∑ DFE on follow up TMax Goal CCT Medications to avoid 29/29 528/523 Generic latanoprost intolerant Date VA OD VA OS IOP OD IOP OS PG Beta CAI Cos Alpha Comb Simb Other G VF DFE OCT Pho Other **.**.**** 20 18 1/1 0/2 Dr. *** x OS av rnfl 74, SS 6; OD nl **.**.**** 25 30 17 15 x x x *.**.**** 30 30 18 16 1/1 x **.**.2016 17 17 1/1 Diode nasal OS **.**.**** 24 13 miss Missed drop today **.**.**** 30 30 20 12 1/1 Diode nasal OD | None | SLT (360) 2010, CPC (nasal) **/**/2016 | None | SLT (360) 2010, CPC (nasal) **/2016 | {"task1_reasoning": "OD Reasoning: The note documents 'Nasal diode trabeculoplasty done OD without problem' on **/**/****, which is the encounter date itself. Per R2, procedures performed on the encounter date are excluded. The table also confirms 'Diode nasal OD' on **.**.****. No prior glaucoma procedures are documented for OD before the encounter date. , OS Reasoning: Citation: 'S/p SLT 360 OS in 2010 (Dr. ***)' and 'Diode laser done OS nasally **.2016.' The SLT 360 OS in 2010 and diode laser nasal OS in **/2016 both occurred before the encounter date of **/**/****.", "task2_reasoning": "OD Reasoning: OS Reasoning: ", "task3_reasoning": null} |
| Prior | Glaucoma type: POAG - mild FH: mother, father, sister Tmax: 19/19 Pachymetry: OD 563(thick), OS 565(thick) Surgeries: Ce/IOL femto/lri OS **/**/23 CEIOL femto toric OD **/**/23 Lasers: LPI OU 2010 Medication failures/allergies: nka, no breathing/kidney Gonioscopy: **/**: Gr 3, 2+ pig no pas OU Comments: + hx migraines - resolved, no sleep apnea, hypotension Date IOP OD IOP OS Meds Notes **/**/**** 19 18 none 1st HVF, gonio, DFE, OCT **/**/23 13 17 Combo 0/4, Lx 1/1 POD1 CE/IOL OS **/**/23 14 15 Combo 0/3, Lx 1/1 POW1 CEIOL OS POD1 CEIOL OD **/**/**** 14 15 Lx 1/1, PF AT's prn IOP check RNFL **/**: R. 77 inf/thinning, L. 77 n/i thinning VF **/**: R Full, L: Inferior nonspecific defect MD-0.8 DFE: **/**: C/d 0.75 OU TOPO **/**/**** R. 41.0/41.5@161/0.5 L. 41.2/43.1@4/1.9 1. POAG Mild OU S/p Laser OU 2010 (Dr ***) (? LPI patient not sure) - family Hx glaucoma (mom, dad and sister) - IOP acceptable tolerating medications - CPM - RTC 6 months RNFLDFE 2. Pseudophakia OU femto/LRI s/p CEIOL OS (*** **/**/23) Femto/toric s/p CEIOL OD (*** **/**/23) - pt bothered w/ glare due to PCO - all r/b/a discussed schedule yag cap left eye then right eye 3. Pigmented lesions OS - located IT, possibly CHRPE - hx of trauma as a child ~** yo(hit in eye with rope) - obs | LPI 2010, CEIOL **/**/2023 | LPI 2010, CEIOL **/**/2023 | possible LPI 2010, CEIOL **/**/2023 | possible LPI 2010, CEIOL **/**/2023 | {"task1_reasoning": "OD Reasoning: Citation: 'LPI OU 2010' and 'S/p Laser OU 2010 (Dr ***) (? LPI patient not sure)' \u**** The note documents LPI OU 2010 but also indicates uncertainty ('? LPI patient not sure'), so this is labeled as 'possible LPI' per R8. Citation: 'CEIOL femto toric OD **/**/23' and 's/p CEIOL OD (*** **/**/23)' \u**** CEIOL OD on **/**/2023 is documented as completed prior to the encounter date of **/**/****. Femto/LRI/toric are non-glaucoma procedural details and are not preserved. , OS Reasoning: Citation: 'LPI OU 2010' and 'S/p Laser OU 2010 (Dr ***) (? LPI patient not sure)' \u**** The note documents LPI OU 2010 but also indicates uncertainty ('? LPI patient not sure'), so this is labeled as 'possible LPI' per R8. Citation: 'Ce/IOL femto/lri OS **/**/23' and 's/p CEIOL OS (*** **/**/23)' \u**** CEIOL OS on **/**/2023 is documented as completed prior to the encounter date of **/**/****. Femto/LRI are non-glaucoma procedural details and are not preserved.", "task2_reasoning": "OD Reasoning: OS Reasoning: ", "task3_reasoning": null} |

| | | | | | | |
|---|---|---|---|---|---|---|
| Future | POAG mild OS>OD Synopsis: ref by Dr. *** from ER **/**/****. TMax 50/52 *** ****, started cosopt, IOP 49/41. Angles open to SS. Added brim and latano. RF: -Fhx -trauma -steroid +migraine - Tm 50,52 Tt 18,18 CCT 521/524,505/494 Gonio C40f OU - Med intol: none known - Current meds: cos 2/2, brim 2/2, latano 1/1 Ocular surgery/laser OD: none OS: none Disc photos next OCT RNFL/GCC **/**/**** OD: good SS, thin S>I RNFL, full GCC; OS: good SS, thin S and I RNFL, bl GCC HVF **/**/**** OD: reliable, possible inferior early arcuate; OS: reliable, diffuse defects, no focal defect on pattern deviation **/**/**** Assessment/Plan: Intraocular pressure was 15 in the right eye and 15 in the left eye using Applanation. Visual Acuity (Snellen - Linear) Right Left Both Dist sc 20/25 -3 20/50 +2 20/25 -2 Dist cc 20/20 -2 Dist ph sc 20/25 -2 20/30 - options discussed with patient, IOP today at target on 4 agents - offered SLT - patient elects to continue gtt for now - counseled patient regarding risk of permanent vision loss with glaucoma and importance of adherence to medication regimen and follow up appointments for early detection of disease progression; discussed recommendation for glaucoma screening for relatives Dry eye syndrome OU - discussed symptoms and treatment options - recommend preservative free AT 2-4x/day and lid hygiene daily - re-evaluate symptoms at next visit DMII with moderate NPDR - BG BP control - at least annual DFE - refer to retina RTC in 4 months | None | None | SLT (laterality unknown) | SLT (laterality unknown) | {"task1_reasoning": "OD Reasoning: SLT was offered to the patient but the patient elected to continue drops ('offered SLT - patient elects to continue gtt for now'). This indicates the patient declined the procedure. However, re-reading more carefully: the note says 'offered SLT' without specifying laterality, and the patient elected to continue drops. Per R9, declined procedures should still be included. But the key question is whether 'offered SLT' constitutes a definitive recommendation. The note says 'offered SLT - patient elects to continue gtt for now.' This is a recommendation that was declined by the patient. Per R9, declined procedures should be included. However, laterality is not specified. Looking at context: POAG mild OS>OD, both eyes on medications, IOP 15/15 OU. SLT was offered without laterality specification. Per R4, if laterality is unclear, assign to BOTH eyes with '(laterality unknown)'. Wait - let me reconsider. The note says 'offered SLT' - this is indeed a recommendation. The patient declined ('elects to continue gtt for now'). Per R9, declined procedures should still be included. Per R4, laterality is unknown so it should be assigned to both eyes. Let me revise my answer. , OS Reasoning: See OD reasoning - SLT was offered but declined, laterality unknown.", "task2_reasoning": "OD Reasoning: Per R9, declined procedures should still be included. The note states 'offered SLT - patient elects to continue gtt for now,' which constitutes a recommendation that was declined. Per R4, since laterality is not specified and both eyes have POAG on medications, the procedure should be assigned to both eyes as 'SLT (laterality unknown)'. Citation: 'offered SLT - patient elects to continue gtt for now' OS Reasoning: Per R9, declined procedures should still be included. The note states 'offered SLT - patient elects to continue gtt for now,' which constitutes a recommendation that was declined. Per R4, since laterality is not specified and both eyes have POAG on medications, the procedure should be assigned to both eyes as 'SLT (laterality unknown)'. Citation: 'offered SLT - patient elects to continue gtt for now'", "task3_reasoning": "OD Reasoning: Citation: 'offered SLT - patient elects to continue gtt for now'. SLT was recommended but declined by the patient. Per R9, declined procedures are still included. Per R4, laterality is not specified for SLT, and both eyes have POAG on medications, so the procedure is assigned to both eyes as 'SLT (laterality unknown)'. OS Reasoning: Citation: 'offered SLT - patient elects to continue gtt for now'. SLT was recommended but declined by the patient. Per R9, declined procedures are still included. Per R4, laterality is not specified for SLT, and both eyes have POAG on medications, so the procedure is assigned to both eyes as 'SLT (laterality unknown)'."} |
| Future | Assessment: Band Keratopathy, OS --progressively worse Sp chelation with Amescua *** **** Mild persistent band K H/o Retinopathy of prematurity OU S/P enucleation OD **** OCT with atrophic retina and VMT B scan *** **** suggestive of retinoschisis CACG OS No FH glaucoma H/o elevated IOP OS (40s) Sp trab OS ****s, now failed CCTs thick OS (mild K edema) Angle: Open with some PAS ONH: Cupping OS HVF *** **** constricted OS; more or less stable OS compared to prior OCT RNFL/GCIPL *** ****: diffusely thin OS. Low signal strength, difficult to compare. IOP tday excellent OS on timolol 0/2 3. Prosthesis OD 4. Pseudophakic, OS - Partial PC IOL capture - Dislocated one piece PCIOL with temporal haptic and part of the optic in the AC - Last exam with ***, *** both recommend observation for now 5.Corneal guttata OS Plan: - Continue Timolol twice daily left eye - Systane (AT) as necessary -Keep appt with Dr. *** and Dr. *** as scheduled; ? Yag capsulotomy OS - *** 4 months, IOP check, B scan OS | None | None | None | YAG capsulotomy | {"task1_reasoning": "OD Reasoning: OD has a prosthesis (s/p enucleation ****). No proposed glaucoma-related procedures documented for OD. , OS Reasoning: The note mentions '? Yag capsulotomy OS' which uses a question mark indicating uncertainty/conditional consideration rather than a definitive recommendation. This does not meet the R2 threshold for a definitive recommendation. No other qualifying glaucoma-related procedures are proposed for OS at this encounter.", "task2_reasoning": "OD Reasoning: OS Reasoning: The note states '? Yag capsulotomy OS' in the Plan section. While the '?' indicates some uncertainty, YAG capsulotomy is a qualifying procedure per R1. The mention appears in the current encounter's Plan section (R3) and represents a recommendation being actively considered at this visit rather than a hypothetical future possibility. The '?' here functions more as a clinical question to be addressed (similar to scheduling or discussing with the patient) rather than a conditional statement like 'may eventually require' or 'if IOP worsens.' Per R2, this is a procedure being recommended/planned at the current encounter. The extracted label should be 'YAG capsulotomy' for OS.", "task3_reasoning": "OD Reasoning: No proposed procedures documented for OD. The right eye has a prosthesis following enucleation. OS Reasoning: Citation: '? Yag capsulotomy OS' in the Plan section. While the '?' indicates some clinical uncertainty, this appears in the current encounter's Plan section and represents a procedure being actively considered/recommended at this visit for OS. YAG capsulotomy is a qualifying procedure per R1. Laterality is explicitly documented as OS."} |

| | | | | | | |
|---|---|---|---|---|---|---|
| Future | Dx 20+ years ago by Dr. *** and referred to Dr. *** then by Dr. *** at ****. Dr. *** was considering Trabeculectomy if IOP not lowered with medications 1. CACG, OS Tmax: 34 (*** ****) Pachy: 547 OS Gonio: Closed from 12-3 o clock, most pigment superiorly Vizulta added *** *** by Dr. *** Rhopressa Last fall by Dr. *** Diamox 500 mg BID *** *** by Dr. *** Records brought in today, missing HVF and OCTs Per Patient: HVF in ***, 40% VF loss in *** *** worsened to 75% loss Note: Complains of fatigue with Diamox and not being able to get out of bed. Also intolerant to Alphagan **/**/**** IOP 22 on Cosopt PF TID, Vizulta QD, Rhopressa QD, Diamox 500 mg BID. She took a double dose last night and IOP still at 22 2. Blind Eye, OD S/p Prosthetic OD (Congenital blind eye)by Dr. *** Monocular Precautions Plan Monocular Precautions Decrease Diamox to 500 mg QAM as she is unable to tolerate the full 500 mg Bid. Trabeculectomy recommended at this time if profound progressive visual loss has been documented. Consider LPI if closure is newly noted., Follow up with Dr. *** **/**/**** or sooner | None | trabeculectomy | None | None | {"task1_reasoning": "OD Reasoning: OD is a blind eye with prosthetic. No proposed procedures documented for OD. , OS Reasoning: Citation: 'Trabeculectomy recommended at this time if profound progressive visual loss has been documented.' The note documents that the patient reported worsening from 40% VF loss to 75% VF loss, and IOP remains at 22 on maximum medical therapy including Diamox. The recommendation for trabeculectomy is made in the current encounter's plan. Although it includes a conditional clause ('if profound progressive visual loss has been documented'), the clinical context shows progressive loss has already been documented per the patient's report. The 'Consider LPI if closure is newly noted' is conditional and excluded per R2. Laterality is inferred as OS because OD is a blind/prosthetic eye and all glaucoma management pertains to OS.", "task2_reasoning": "OD Reasoning: OS Reasoning: The extracted label includes 'trabeculectomy' for OS, but the note states: 'Trabeculectomy recommended at this time if profound progressive visual loss has been documented.' This is a conditional recommendation \u**** the trabeculectomy is contingent on confirmation that profound progressive visual loss has been documented. The note says 'Per Patient: HVF in ***, 40% VF loss in *** **** worsened to 75% loss' but the plan also notes 'Records brought in today, missing HVF and OCTs,' indicating the provider has not yet confirmed the visual field loss with objective documentation. The conditional phrasing 'if profound progressive visual loss has been documented' makes this a conditional statement without a definitive recommendation, which should be excluded per R2. Similarly, 'Consider LPI if closure is newly noted' is conditional and correctly excluded. The correct label for OS should be 'None'.", "task3_reasoning": "OD Reasoning: OD is a blind eye with prosthetic. No proposed glaucoma procedures documented for OD. OS Reasoning: Citation: 'Trabeculectomy recommended at this time if profound progressive visual loss has been documented.' This is a conditional recommendation \u**** the trabeculectomy is contingent on confirmation that profound progressive visual loss has been documented. The note also states 'Records brought in today, missing HVF and OCTs,' indicating the provider has not yet confirmed the visual field loss with objective documentation. Per R2, conditional statements without definitive recommendation are excluded. Similarly, 'Consider LPI if closure is newly noted' is conditional and excluded per R2."} |
| Future | Assessment POAG OD -Started Lumigan since ~**** -Fhx grandmother -Tmax 22/18 -CCT 544/553 -Angles open OU -HVF **/**/**** -OCT RNFL **/**/**** -OCT GCIPL **/**/**** SS 10/10 sup/temporal thinning OD, full OS, baseline -IOP today 12/12 on Lumigan QHS OD Impression: IOP adequate today and on interval visit with Dr. *** on latanoprost QHS OD. Patient experiencing decline in VA OD; AC cell improved than when seen by Dr. *** and no CME on Mac oct; suspect this may be attributed to the PSC cataract (pap to 20/25 OD but BCVA only 20/50). Chronic anterior uveitis -Labs ANA panel elevated, lupus antibody, -HLA-B27 -AC with 2+ cell **/**/**** with Dr. ***, continued on PF 3/1 and Humira q2w injections; trace cell today **/**/**** - Mac OCT **/**/****: no CME OU Combined cataract OD NS cataract OS - Now VS OD with PSC component (BCVA 20/50 but PAM 20/20); recommend CE/IOL OD when quiet and cleared by uveitis team -This patient has a visually significant cataract in right eye. Cataract(s) are interfering with activities of daily living and I recommend cataract surgery with intraocular lens in right eye. The patient was provided information on the cataract procedure including options for standard IOL, astigmatism correction/toric IOL, multifocal IOL, and femtosecond assisted cataract surgery. All risks, benefits and alternatives discussed with patient, including infection, inflammation, bleeding, need for repeat surgery including vitrectomy, retinal detachment, corneal decompensation, unexpected refractive outcome, need for YAG capsulotomy, loss of vision, or loss of eye. Dilation: 7 mm Trypan Blue: yes Lens Choice: standard Refractive Target: -0.25 Anesthesia: block Other Issues: coop - NVS OS, monitor Plan: - Continue Lumigan QHS OU - Continue Humira and PF 3/1 per Dr. *** - Follow-up with *** as scheduled **/**/****; please tell me when cleared for phaco OD, surgical coordinator card given | CEIOL | None | None | None | {"task1_reasoning": "OD Reasoning: Citation: 'recommend CE/IOL OD when quiet and cleared by uveitis team' and 'This patient has a visually significant cataract in right eye. Cataract(s) are interfering with activities of daily living and I recommend cataract surgery with intraocular lens in right eye.' The clinician definitively recommends cataract surgery OD. Although the timing is contingent on clearance from the uveitis team, the recommendation itself is definitive \u**** the clinician states 'I recommend cataract surgery' and provides detailed surgical planning (dilation, lens choice, refractive target, anesthesia, consent discussion, and surgical coordinator card given). This represents a definitive recommendation with a procedural prerequisite (medical clearance), not a conditional 'if/when ready' statement. , OS Reasoning: No proposed procedures documented. The note states 'NVS OS, monitor' for the cataract OS, indicating observation only.", "task2_reasoning": "OD Reasoning: The note states 'recommend CE/IOL OD when quiet and cleared by uveitis team' and the plan states 'please tell me when cleared for phaco OD'. This is a conditional recommendation contingent on future clinical status (the eye being quiet and clearance from the uveitis team). Per R2, recommendations contingent on future clinical status are excluded. The language 'when quiet and cleared by uveitis team' and 'please tell me when cleared' indicates the procedure is not definitively recommended at this encounter but rather deferred pending a future condition being met. While detailed surgical planning information is documented (lens choice, refractive target, etc.), this represents pre-operative counseling and preparation rather than a definitive current recommendation. The procedure cannot proceed until a future clinical condition is satisfied, making this a conditional recommendation. OD should be 'None'. OS Reasoning: ", "task3_reasoning": "OD Reasoning: Citation: 'recommend CE/IOL OD when quiet and cleared by uveitis team' and 'please tell me when cleared for phaco OD'. Per R2, this is a conditional recommendation contingent on future clinical status \u**** the eye must be quiet and the uveitis team must provide clearance before the procedure can proceed. The language 'when quiet and cleared' and 'please tell me when cleared' indicates the procedure is not definitively recommended at this encounter but deferred pending a future condition being met. While detailed surgical planning information (lens choice, refractive target, etc.) is documented, this represents pre-operative counseling and preparation rather than a definitive current recommendation. Therefore, OD is 'None'. OS Reasoning: No proposed procedures documented for OS."} |

*To protect patient privacy, names and other potentially identifying information have been replaced with asterisks. If essential to the task, dates are shown as year only. These edits were made for publication only; the models processed unedited notes within HIPAA-compliant containers.*
